# Tajima *D* test accurately forecasts Omicron / COVID-19 outbreak

**DOI:** 10.1101/2021.12.02.21267185

**Authors:** Ting-Yu Yeh, Gregory P. Contreras

**Author notes:** Ting-Yu Yeh is Corresponding author.

## Abstract

On 26 November 2021, the World Health Organization designated the SARS-CoV-2 variant B.1.1.529, Omicron, a variant of concern. However, the phylogenetic and evolutionary dynamics of this variant remain unclear. An analysis of the 131 Omicron variant sequences from November 9 to November 28, 2021 reveals that variants have diverged into at least 6 major subgroups. 86.3% of the cases have an insertion at amino acid 214 (INS214EPE) of the spike protein. Neutrality analysis of *DH* (−2.814, *p*<0.001) and Zeng’s *E* (0.0583, *p*=1.0) tests suggested that directional selection was the major driving force of Omicron variant evolution. The synonymous (*D*_syn_) and nonsynonymous (*D*_nonsyn_) polymorphisms of the Omicron variant spike gene were estimated with Tajima’s *D* statistic to eliminate homogenous effects. Both *D* ratio (*D*_nonsyn_/*D*_syn_, 1.57) and Δ*D* (*D*_syn_-*D*_nonsyn_, 0.63) indicate that purifying selection operates at present. The low nucleotide diversity (0.00008) and Tajima *D* value (−2.709, *p*<0.001) also confirms that Omicron variants had already spread in human population for more than the 6 weeks than has been reported. These results, along with our previous analysis of Delta and Lambda variants, also supports the validity of the Tajima’s *D* test score, with a threshold value as −2.50, as an accurate predictor of new COVID-19 outbreaks.

## Introduction

On 26 November 2021, World Health Organization designated the SARS-CoV-2 variant B.1.1.529, Omicron, a variant of concern based on its unique mutations, and unusual features suggesting that therapeutic monoclonal antibodies, such as Regeneron, may be less effective against Omicron. Evidences that new mutations in Omicron could have an impact on viral transmission or the severity of illness are still under investigation (World Health Organization, 2021). Omicron variants were first reported in the Gauteng province, South Africa on November 9, 2021, and shortly it was detected in recent travelers to Belgium, Botswana, Canada, Hong Kong, Austria, Australia, Portugal, Israel, United Kingdom, Netherland, and the United States. One of the major concerns is that Omicron variants contain more than 30 mutations to the spike protein (A67V, Δ69-70, T95I, G142D/Δ143-145, Δ211/L212I, ins214EPE, G339D, S371L, S373P, S375F, K417N, N440K, G446S, S477N, T478K, E484A, Q493R, G496S, Q498R, N501Y, Y505H, T547K, D614G, H655Y, N679K, P681H, N764K, D796Y, N856K, Q954H, N969K, L981F). Some of these changes have been previously identified in Delta or Alpha variants and are linked to heightened infectivity and the ability to evade infection-blocking antibodies (Callaway, 2021). The likelihood of higher transmission rates has led multiple countries to respond quickly to the Omicron variant.

The rapid increase in Omicron variant cases was found in the Gauteng province in November, particularly in schools and among young people. Preliminary evidence from genotyping tests suggests that Omicron may have been in circulation for quite some time in South Africa (Callaway, 2021). To date, phylogenetic and evolutionary status of Omicron variants are still not clear. Here we analyze the phylogenetic relationship and selection pressure among the 131 available sequences of Omicron variants.

## Materials and Methods

131 complete SARS-CoV-2 genome sequences excluding low coverage from November 9 to November 28, 2021 in 9 countries: Austria (*N*=1), Australia (*N*=1), Belgium (*N*=1), Canada (*N*=1), Botswana (*N*=17), Hong Kong, China (*N*=2), Italy (*N*=1), South Africa (N=105), and United Kingdom (UK, *N*=2) were collected from the Global Initiative on Sharing All Influenza Data (GISAID) (https://www.gisaid.org/). Sequence data in this study is available and deposited at Figshare website (10.6084/m9.figshare.17105090).

FASTA files of viral sequences were downloaded and first aligned using MAFFT 7 software (Katoh and Standley, 2013, Kuraku et al, 2013). Phylogenetic relationships between Omicron variants were analyzed using the neighbour-joining method and Jukes-Cantor substitution model with bootstrap resampling number set as five. The radial phylogenetic tree was generated by exporting the tree file in Newick format by MAFFT. The FigTree software (version 1.4.2) was used to display the cladogram (Rambaut, 2021).

The polymorphisms of the SARS-CoV-2 Omicron variants was analyzed based on the site-frequency spectrum: (1) Tajima’s *D* test, (2) normalized Wu and Fay’s *DH* test, and (3) Zeng’s *E* test, using DNASP6 software with the bat SARS-CoV-2 WIV04 (GenBank MN996528.1) as the outgroup sequence (Rozas, et al., 2017). Statistical significance of observed values of different tests was obtained by coalescent simulation with intermediate recombination after 10,000 replicates. The probability for each statistic was calculated as the frequency of replicates with a value lower than the observed statistic (two-tailed test) by DNASP6 (Rozas, et al., 2017).

A modified Tajima’s *D* statistic was used to examine purifying selection based on non-synonymous (*D*_nonsyn_) versus synonymous sites (*D*_syn_) of SARS-CoV-2 genes was calculated as previously described (Hahn et al, 2002, Hughes, et al., 2005, Yeh and Contreras, 2021). The average number of pairwise synonymous differences (*k*S) and the average number of nonsynonymous pairwise differences (*k*N), the number of synonymous segregating sites (S_S_), and the number of nonsynonymous segregating sites (S_N_) were computed with equation: (1) S*_S_=S_S_/a1, (2) S*_N_=S_N_/a1, where a1 is defined as 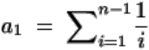 (Tajima, 1989, Hughes, et al., 2005). *D*_syn_ was defined as *k*S - S*_S_, divided by the standard error of that difference, and *D*_nonsyn_ was defined as *k*N - S*_N_, divided by the standard error of that difference (Tajima, 1989, Hughes, et al., 2005).

## Results and Discussion

### The insertion mutation Ins214EPE at the spike protein is present in some Omicron variants

The phylogenetic tree identifies at least 6 major subgroups of 131 Omicron variant sequences after rooting with an outgroup virus sequence of SARS-CoV-2 WIV04 from Wuhan, China (Figure 1). 113 Omicron cases (86.3%) contain an insertion of nine nucleotides (GAGCCAGAA) between nucleotide 22204 and 22205 according to WIV04 sequence (Figure 2). This generates an insertion of three amino acids (Glutamic acid-Proline-Glutamic acid) (INS214EPE) in the N-terminal domain (NTD) of the spike protein.

**Figure 1.**
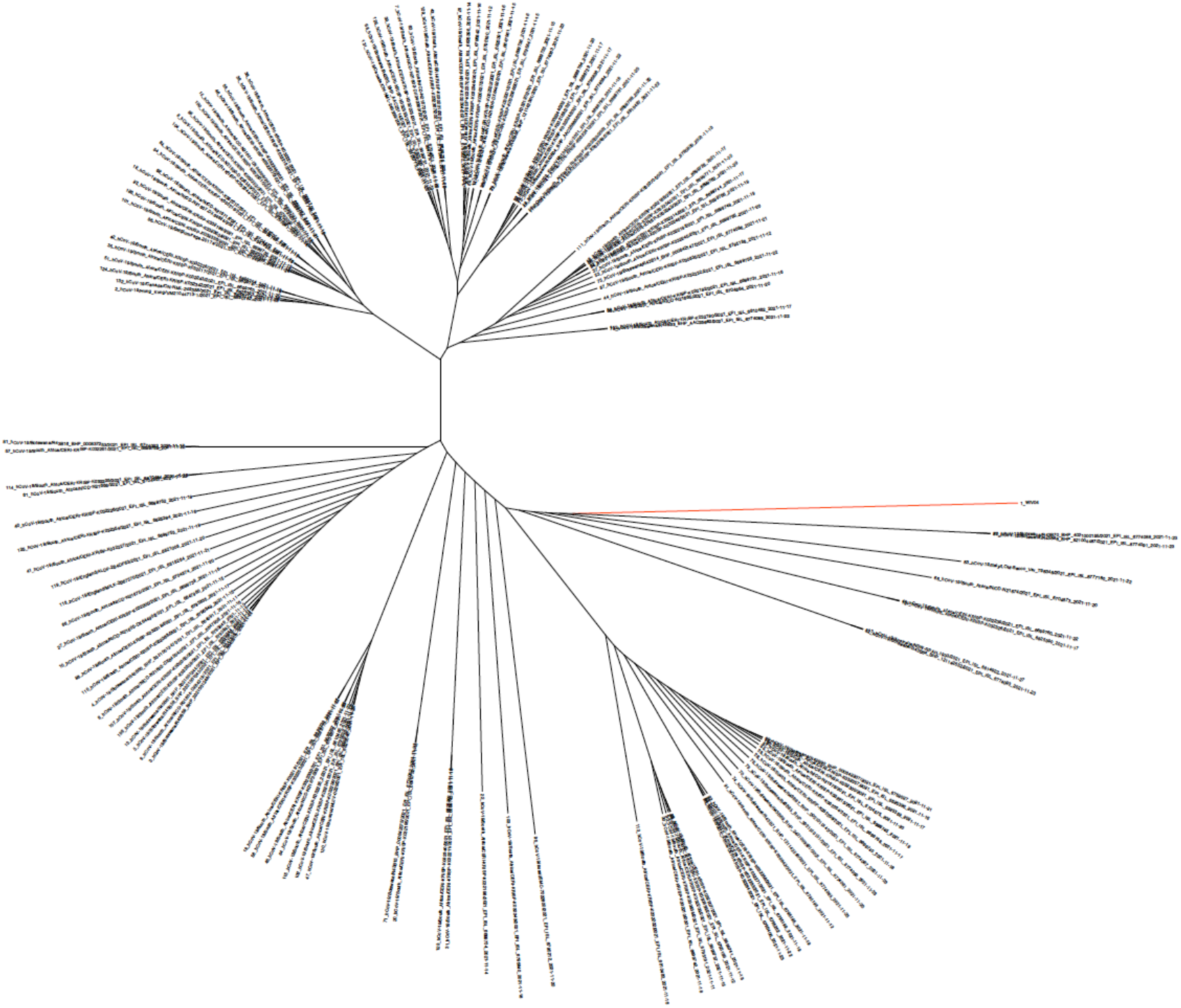
Rooted phylogenetic tree of SARS-CoV-2 genomes of Omicron variants, November 9 to November 28, 2021. Alignments of viral sequences were generated using MAFFT, and the phylogenetic tree was visualized using FigTree. Rooting was done by introducing SARS-CoV-2 WIV04 (red line, accession number MN996528) as an outgroup virus.

**Figure 2.**
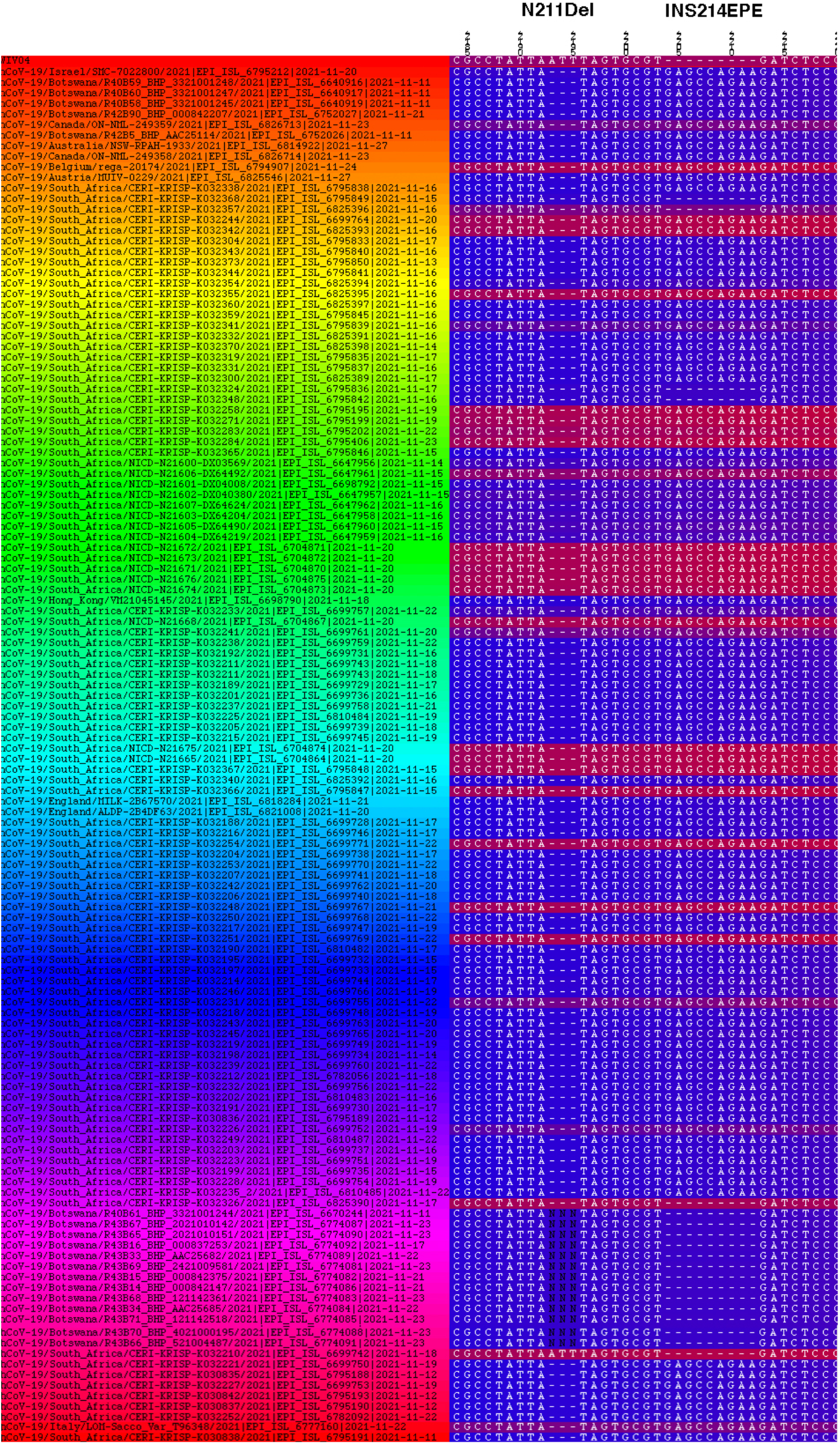
Alignment of Omicron variant with and without insertion at the amino acid 214 of the spike protein.

Resende et al. has reported that most ins214 motifs were rare in sequences of different lineages of SARS-CoV-2 (A.2.4, B.1, B.1.1.7, B.1.177, B.1.2, B.1.214, and B.1.429) with an insertion motif of 3 or 4 amino acids (AKKN, KLGB, AQER, AAG, KFH, KRI, and TDR) (Resende, et al, 2021). Interestingly, the ins214 with four amino acids (ins214GATP, ins214GATP, ins214GATS) were also present in bat SC2r-CoV isolated in China (RmYN02), Thailand (RacCS203), and Japan (Rc-o319), respectively. Although amino acid identity at ins214 vary among SARS-CoV-2 or SC2r-CoV lineages, the insertion size (3 or 4 amino acids) is also conserved in Omicron. It suggested that this region of NTD is susceptible to a mutation or insertion. Whether or not INS214EPE affects spike protein function or immune response requires further investigation.

### Tajima’s *D* test is able to forecast new Omicron outbreaks

We have previously used neutrality tests to analyze the selection pressure of SARS-CoV-2 in the shipboard quarantine on the Diamond Princess (Yeh and Contreras, 2021a) and the influence of full vaccination coverage on Delta variants among different countries (Yeh and Contreras, 2021b). We also proposed that Tajima’s *D* test, a popular statistic in population and evolution genetics, can provide a promising tool to forecast new COVID-19 outbreaks (Yeh and Contreras, 2021b).

Here the selection pressure of Omicron variants was first analyzed by multiple neutrality tests. Within 131 Omicron variant sequences, 75 mutation sites were identified with the nucleotide diversity (π, the average number of nucleotide differences per site between two sequences) equal to 0.00008, which is significantly lower than earlier outbreak of Delta variants in UK (0.0004-0.0006, *N*=376, March 26 to April 22, 2021), India (0.0004-0.0006, *N*=67, January 1 to March 11, 2021), or Australia (0.0006, *N*=75, April 9 to May 6, 2021) (Yeh and Contreras, 2021b). Tajima *D* test was calculated to compare π and total polymorphism (Tajima, 1989). Tajima’s *D* values were negative and significantly deviated from zero (−2.709, *P* < 0.001) among the whole genome sequences of Omicron variants. The negative values of Tajima *D* were also detected in the ORF1ab (−2.617, *P* < 0.001) and the spike gene (−1.948, *P* < 0.005). This result indicates an excess of nucleotide variants of low frequency (Table 1), and strong selection and/or demographic expansion was operating in Omicron.

**Table 1.**
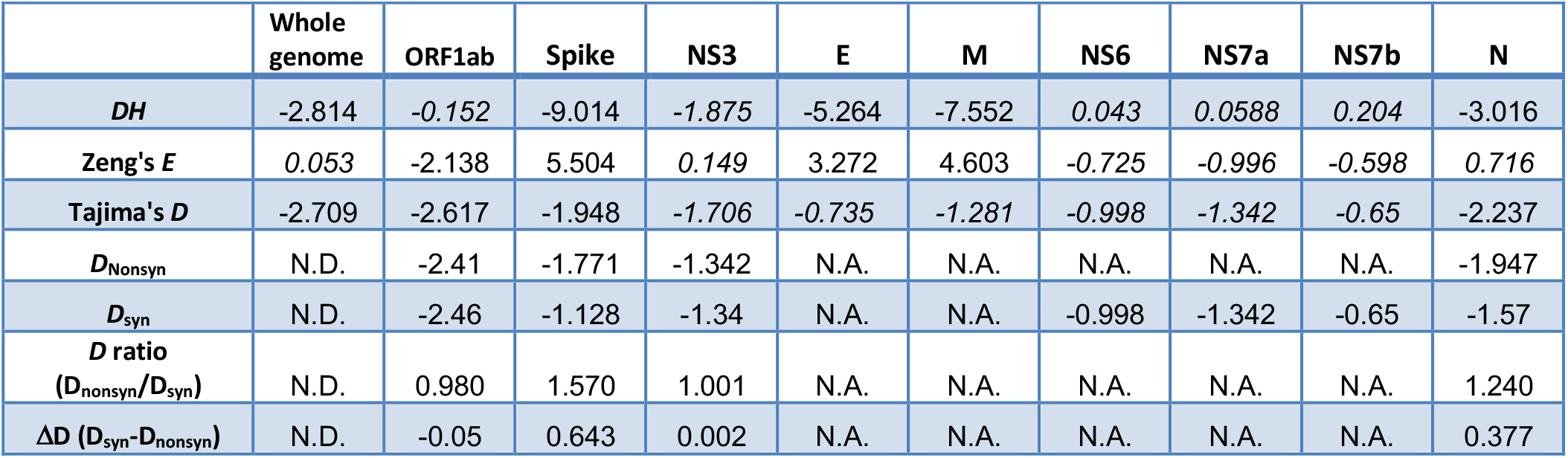
Neutrality analysis of SARS-CoV-2 Omicron variants of the full genome and the viral genes. The statistical significance was estimated using 10,000 coalescent simulations in DNASP6. Not significant values (*P*>0.05) are indicated in italic. NS8 is excluded in this table because no mutations were detected. (N.D., not determined; N.A., not applicable).

We have previously shown that Tajima *D* values decreased as SARS-CoV-2 Delta and lambda variants spread in human populations. One to three weeks after Tajima *D* fell below −2.50, Delta variant outbreaks emerged in India and the UK (Yeh and Contreras, 2021b). Taken together, the low π and Tajima *D* values suggested that Omicron variants have most likely spread within a population weeks before the samples were collected. **This data also agreed with our previous proposal that Tajima’s *D* test is useful to forecast new COVID-19 outbreaks regardless of the sample sizes of different variants** (Yeh and Contreras, 2021b).

### Purifying selection was operating in Omicron variants

Application of Tajima *D* is limited by the difficulty in distinguishing the influence of both selection pressure and demographic expansion. To overcome this problem, we included normalized *DH* test and Zeng’s *E* test in our analysis (Zeng et al., 2006, Yeh and Contreras, 2021b). The normalized *DH* test is affected by directional selection but insensitive to demographic expansion. Zeng’s *E* test is very sensitive to population growth immediately after a sweep (Zeng et al., 2006). Genomic polymorphisms of Omicron variants showed significantly negative *DH* values (−2.814, *P*<0.05), but *E* values were not significantly different from zero (0.0583, *P*>0.1) (Table 1). The results were similar after confining our analysis to the spike gene (*DH*, −9.014, *p*<0.001; Zeng’s *E*, 5.504, *p*<0.001) (Table 1), suggesting that directional selection was the major driving force, without significant influence by the demographic expansion.

We and others have shown the limitations to application of the inter-species divergence, the dN/dS (ω) test, for analyzing selection pressure of SARS-CoV-2 previously (Mugal, et al., 2014, Kang et al., 2021, Yeh and Contreras, 2021a, Yeh and Contreras 2021b). Therefore, here we determined the purifying selection using a modified Tajima’s *D* statistics instead of dN/dS (ω) test. Under purifying selection, the frequency distribution of non-synonymous polymorphisms is negatively skewed relative to the distribution of synonymous polymorphisms. Therefore, it takes more negative values for non-synonymous (*D*_nonsyn_) than for synonymous sites (*D*_syn_) of a given gene (Hahn et al, 2002, Hughes, et al., 2005, Yeh and Contreras, 2021b). One of the major advantages of *D*_nonsyn_ and *D*_syn_ analysis is that it is independent of sample size, which allows us to compare *D*_nonsyn_ and *D*_syn_ values among different data sets (Hughes et al., 2008). The same excess of low-frequency alleles in non-synonymous polymorphism was also shown by the *D*_nonsyn_ values of the spike (−1.771, *p*<0.001) and N gene (−1.943,, *p*<0.005) (Table 1). **We conclude that purifying selection led to constraints on the neutral mutations at non-synonymous sites of the spike gene of Omicron variants**.

Negative values of Tajima *D* could be caused by a bottleneck event rather than selection, but the bottleneck effect should affect all types of polymorphism equally (Tajima, 1989). Hahn et al. have reported that *D*_nonsyn_ is disproportionally lower than *D*_syn,_ because non-synonymous and synonymous mutations are affected unequally by purifying selection (Hahn et al., 2002). The value of Δ*D* (*D*_syn_-*D*_nonsyn_) increased as purifying selection became stronger, and Δ*D* can eliminate the homogenous effects (demographic expansion, selective sweep etc.). Δ*D* values of the spike and N gene of SARS-CoV-2 Omicron variants are positive (0.643 and 0.377, *p*<0.005) (Table 1). Austin Hughes showed that the standard error terms in both equations of *D*_nonsyn_ and *D*_syn_ cancel out the sample size effect when *D* ratio (*D*_nonsyn_/*D*_syn_) was applied to the data analysis (Hughes, 2008). ***D* ratio values of the spike and N gene were significantly more than 1 (1.57 and 1.24, Table 1), suggesting that purifying selection pressure of Omicron spike and N gene was operating**. The values of Δ*D* and *D* ratio of ORF1ab (−0.05 and 0.980, *p*<0.001) were insignificant, consistent with the idea that the spike and N gene have been under more selective constraint than ORF1ab (Chaw et al., 2020). So far, no evidence has shown that vaccination or other mitigation procedures causes positive selection of Omicron variants.

### Perspectives

Previously we found that one to three weeks after the *D* value fell below −2.50, Delta outbreaks emerged in India and UK, and the lambda variant outbreak in south America (Yeh and Contreras, 2021b). This led to our proposal that Tajima *D* test with a cut-off threshold value as −2.50, can predict new SARS-CoV-2 outbreak (Yeh and Contreras, 2021b). Despites of the small sequence sample size (131 samples) in this study, we detect a strong negative value of Tajima *D* in Omicron variants. This finding also confirmed that the Omicron outbreak had emerged sometime before the current breakout as the Tajima *D* test is sensitive to detect it and an efficient predictor of future outbreaks. This study demonstrates that rapid genomic sequence surveillance is essential, and Tajima *D’* tests should be included to forecast future outbreaks in different geographic populations.

## Data Availability

All data produced are available online at Figshare website (10.6084/m9.figshare.17105090).

https://10.6084/m9.figshare.17105090

## Disclosure statement

No potential conflict of interest was reported by the author(s).

## Author contributions

All authors contributed to study concept, rationale, and initial manuscript drafts, interpretation of data, and final manuscript preparation. All authors have read and approved the final version of the manuscript.

## Funding

No funding

## Ethical approval

None declared.

## Notes

### Competing Interest Statement

The authors have declared no competing interest.

### Funding Statement

This study did not receive any funding

## References

Callaway, E. (2021) Heavily mutated Omicron variant puts scientists on alert. Nature. https://doi.org/10.1038/d41586-021-03552-w

Chaw, S.M., Tai, J.H., Chen, S.L., Hsieh, C.H., Chang, S.Y., Yeh, S.H., Yang, W.S., Chen, P.J., Wang, H.Y. (2020) The origin and underlying driving forces of the SARS-CoV-2 outbreak. J Biomed Sci., 27, 73.

Katoh, K, Standley, D.M. (2013) MAFFT multiple sequence alignment software version 7: improvements in performance and usability. Mol Biol Evol. 30, 772–80.

Kuraku S, Zmasek CM, Nishimura O, Katoh K. (2013) aLeaves facilitates on-demand exploration of metazoan gene family trees on MAFFT sequence alignment server with enhanced interactivity. Nucleic Acids Res.,41, W22–8.

Hahn, M.W., Rausher, M.D., Cunningham, C.W. (2002) Distinguishing between selection and population expansion in an experimental lineage of bacteriophage T7. Genetics, 161, 11–20.

Hughes, A.L., (2005) Evidence for Abundant Slightly Deleterious Polymorphisms in Bacterial Populations. Genetics, 169, 533–538.

Hughes, A.L., Friedman, R., Rivailler, P., French, J.O., (2008) Synonymous and nonsynonymous polymorphisms versus divergences in bacterial genomes. Mol Biol Evol., 25, 2199–2209.

Kang, L., He, G., Sharp, A.K., Wang, X., Brown, A.M., Michalak. P., Weger-Lucarelli, J. (2021) A selective sweep in the Spike gene has driven SARS-CoV-2 human adaptation. Cell, 184, 4392–4400.

Mugal, C.F., Wolf, J.B., Kaj, I. (2013) Why time matters: codon evolution and the temporal dynamics of dN/dS. Mol Biol Evol., 31, 212–231.

Resende, P. C., Naveca, F. C., Lins, R. D. et al. (2021) The ongoing evolution of variants of concern and interest of SARS-CoV-2 in Brazil revealed by convergent indels in the amino (N)-terminal domain of the Spike protein. DOI: 10.1101/2021.03.19.21253946

Rozas, J., Ferrer-Mata, A., Sánchez-DelBarrio, J.C., Guirao-Rico, S., Librado, P., Ramos-Onsins, S.E., Sánchez-Gracia, A. (2017) DnaSP 6: DNA Sequence Polymorphism Analysis of Large Data Sets. Mol Biol Evol., 34, 3299–3302.

Rambaut A. FigTree, version 1.4.2. San Fransisco: GitHub, Inc.; 2021. Available from: https://github.com/rambaut/figtree/releases

Tajima, F. (1989) Statistical method for testing the neutral mutation hypothesis by DNA polymorphism. Genetics, 123, 585–595.

World Health Organization. (2021) Update on Omicron. https://www.who.int/news/item/28-11-2021-update-on-omicron.

Yeh, T.Y., Contreras, G.P. (2020) Emerging viral mutants in Australia suggest RNA recombination event in the SARS-CoV-2 genome. Med J Aust, 213, 44–44.e1.

Yeh, T.Y., Contreras, G.P. (2021a) Viral transmission and evolution dynamics of SARS-CoV-2 in shipboard quarantine. Bull World Health Organ., 99, 486–495.

Yeh, T.Y., Contreras, G.P. (2021b) Full vaccination is imperative to suppress SARS-CoV-2 Delta variant mutation frequency. MedRxiv, https://doi.org/10.1101/2021.08.08.21261768

Zeng, K., Fu, Y.X., Shi, S., Wu, C.I. (2006) Statistical tests for detecting positive selection by utilizing high-frequency variants. Genetics, 174, 1431–1439.

